# Clinical properties of the Short Mood and Feelings Questionnaire: Development of a free calculator based on a Brazilian High-Risk Cohort Study

**DOI:** 10.1101/2025.02.17.25322370

**Authors:** Gabriele dos Santos Jobim, João Villanova do Amaral, João Pedro Gonçalves Pacheco, Ary Gadelha, Euripedes Constantino Miguel, Rodrigo Affonseca Bressan, Pedro Mario Pan, Luis Augusto Rohde, Giovanni Abrahão Salum, Mauricio Scopel Hoffmann

**Author notes:** **Corresponding author:** Dr. Mauricio Scopel Hoffmann. Universidade Federal de Santa Maria, Avenida Roraima 1000, building 26, office 1353, Santa Maria, 97105-900, Brazil (UFSM). I declare that I have full access to all the data in the study and had final responsibility for the decision to submit it for publication.

## Abstract

**Background:** The Short Mood and Feelings Questionnaire (SMFQ) is a validated tool for assessing depressive symptoms in youth, though no specific cut-point exists for the Brazilian population. Item response theory (IRT) and interval likelihood ratios (ILRs) offer refined methods to monitor symptoms but involve complex calculations that hinder clinical implementation.

**Methods:** Cross-sectional data were drawn from an urban school-based sample (Brazilian High-Risk Cohort Study in 2018-2019, n=1,905, aged 14-23, 46.6% females). Diagnoses were based on Development and Well-Being Assessment (DAWBA) clinical ratings. SMFQ factor scores were estimated using IRT and transformed into T-scores. ROC curves evaluated diagnostic properties for internalizing- and externalizing-spectrum disorders. A calculator was developed to estimate post-test probabilities from T-scores using ILRs. Sensitivity analysis excluded MDD as a comorbid diagnosis.

**Results:** ROC curve analyses suggested a sum score cut-off of >6 and a T-score of >55 for detecting MDD. The SMFQ showed good accuracy for internalizing conditions (AUC > 0.8) but low for attention and externalizing disorders (AUC < 0.7). ILRs for internalizing conditions ranged from 0.12 (95% CI: 0.07–0.19) to 29.98 (95% CI: 11.99–75), with post-test probabilities exceeding pre-test probabilities for scores above the cut-off. Sensitivity analysis confirmed findings when excluding MDD. Including ILRs significantly improved predictive models over dichotomous cut-offs.

**Conclusion:** The application of ILRs based on IRT T-scores improved SMFQ’s predictive ability for internalizing-spectrum conditions, regardless of comorbidity. A calculator can integrate these methods into clinical practice, supporting real-time data-driven decisions.

## Introduction

Major depressive disorder (MDD) is a leading cause of disability, particularly among young people (Marx et al., 2023; Vos et al., 2020). Its detection remains neglected, especially in low- and middle-income countries (LMICs) (Collins et al., 2011; Kieling et al., 2011), despite efforts to improve screening (US Preventive Services Task Force et al., 2023). The Mood and Feelings Questionnaire (MFQ) (Angold et al., 1995; Costello & Angold, 1988; Burleson Daviss et al., 2006; Wood et al., 1995) and its brief version, the Short MFQ (SMFQ) (Angold & Costello, 1987; Thabrew et al., 2018; Thapar & McGuffin, 1998; Turner et al., 2014; Eyre et al., 2021), are widely validated scales to assess MDD symptoms in youth. While the MFQ shows equivalence across gender and ethnicity (Banh et al., 2012; Wood et al., 1995; Burleson Daviss et al., 2006), literature reports varying thresholds across settings (Thabrew et al., 2018; Thapar & McGuffin, 1998; Turner et al., 2014). Although the SMFQ has been validated in Brazilian Portuguese (Pinto, 2014; Sucupira et al., 2017), no specific cut-off exists for this population.

Beyond adapting cut-offs, alternative scoring methods are needed. While traditional sum scores assume equal symptom weights (e.g. feeling miserable is equivalent to feeling restless), Item Response Theory (IRT) accounts for item-specific properties, generating scores that can be converted into a common metric like T-scores (Cappelleri et al., 2014; de Beurs et al., 2022). However, sum scores are manually computable, while IRT-based scores require computation, limiting clinical use. Web-based tools can facilitate IRT adoption and enhance symptom monitoring, particularly in LMICs, where MDD detection remains low (Collins et al., 2011; Kieling et al., 2011).

Likelihood ratios link test results to the odds of a condition by comparing their probability in affected and unaffected individuals (Grimes & Schulz, 2005; Gallagher, 1998). Typically, they are derived by dichotomizing continuous data, yielding positive and negative likelihood ratios (Hayden & Brown, 1999). However, this assumes all results above the threshold equally increase risk, leading to information loss. Interval likelihood ratios (ILRs), also called stratum-specific or multilevel likelihood ratios, address this by capturing varying test properties across score ranges, refining post-test probability estimates (Brown & Reeves, 2003; Grimes & Schulz, 2005; Peirce & Cornell, 1993; Simel et al., 1993). Despite their value, ILRs remain underexplored in psychiatry (Schmitz et al., 2000) and untested with MFQ or SMFQ scales. Like IRT, their adoption is hindered by computational demands.

Depression screening tools assess symptoms that are largely transdiagnostic across nosological systems like the DSM-5 (Forbes et al., 2024). Moreover, MDD frequently coexists with other internalizing and externalizing conditions (McGrath et al., 2020; Caspi et al., 2020). Therefore, it is possible that while screening for depression, clinicians can also detect other conditions. While the SMFQ effectively identifies clinical depression, its transdiagnostic potential remains unexplored. Investigating this could broaden its applicability and optimize care by reducing reliance on multiple tools.

Therefore, the present study aims to estimate 1) IRT-based SMFQ T-scores for a urban school-based sample of young people in Brazil, 2) determine its cut-off points for MDD and 3) for various mental health conditions, including Generalized Anxiety Disorder (GAD), Social Anxiety Disorder (SAD), Panic Disorder or Agoraphobia (PD/AG), Post-Traumatic Stress Disorder (PTSD), Attention-Deficit/Hyperactivity Disorder (ADHD), and Conduct Disorder (CD), as well as 4) generate a calculator to estimate post-test probabilities based on ILR of SMFQ scores. As depressive symptoms are pervasive across many diagnostic categories, we hypothesize that SMFQ will present similar ROC curves, epidemiological properties and likelihood ratios to detect different mental health conditions.

## Methods

### Study design and participants

This study analyzed data from the 8-year follow-up (2018–2019) of the Brazilian High Risk Cohort Study for the Development of Childhood Psychiatric Disorders (BHRCS) (Salum et al., 2015). Initiated in 2010, the cohort recruited 2,511 children (ages 6–12) from public schools in two Brazilian capitals (Porto Alegre and São Paulo). Trained interviewers collected household parent reports, while trained psychologists evaluated children. Participants were classified into random or high-risk subsamples based on early psychiatric symptoms or high family risk for psychopathology. The 8-year follow-up included 1,905 participants (79.3% retention), ages 14–23. For this analysis, inclusion criteria were having responded to the SMFQ (n=1633).

This study was approved by the ethics committee of the University of São Paulo [IORG0004884/National Council of Health Registry number (CONEP): 15.457/Project IRB registration number: 1132/08] and followed the ethical standards of the 1964 Declaration of Helsinki and its amendments or comparable ethical standards. Participation was voluntary, with written consent and assent obtained from caregivers and youths, respectively.

### Measures

#### Development and Well-being Assessment (DAWBA)

To assess mental health conditions, the Brazilian-Portuguese version of the Development and Well-being Assessment (DAWBA) was administered to parents by a lay interviewer and to participants by psychologists (Goodman et al., 2000). This semi-structured interview relates to DSM-5 criteria, generating diagnostic probabilities. Certified child psychiatrists evaluated these probabilities and rated responses to determine diagnoses. Conditions with prevalence ≥ 2% in this sample were analyzed: MDD, GAD, SAD, PD/AG, PTSD, ADHD and CD, as well as a group of internalizing conditions (all but ADHD and CD) and any condition.

#### Short Mood and Feelings Questionnaire (SMFQ)

The SMFQ is a 13-item measure used to assess depression symptoms over the past two weeks in the young (Angold et al., 1995). It is highly correlated to the MFQ but requires half the completion time (Angold et al., 1995; Costello & Angold, 1988). Participants completed the self-report version at the BHRCS 8-year follow-up. Items are rated on a 3-point Likert scale (0 = not true, 1 = sometimes, 2 = true), yielding a total score of 0–26. The authors recommend a clinical cut-off of ≥ 8 for children and adolescents (Angold et al., 1995). This measure has been translated and validated to Brazilian Portuguese (Pinto, 2014; Sucupira et al., 2017). In this analysis, we used an unidimensional model for IRT (Messer et al., 1995; Sharp et al., 2006).

### Statistical analysis

To enhance the generalizability, we utilized sampling weights and inverse probability weight (IPW) calculated by Villanova et al. (2025, Preprint). Sampling weight adjusts for the high-risk stratification used in the BHRCS using propensity score (Martel et al., 2017), while IPW uses predictive variables for follow-up response to address sample attrition. The product of these two weights was used in this study. Mean SMFQ sum scores were compared between those with and without DAWBA diagnoses for conditions with prevalence ≥ 2% in this sample.

#### Item Response Theory

IRT analyses evaluated the SMFQ psychometric properties. A graded response model (GRM) fit the SMFQ items in a unidimensional structure (Samejima, 1997; Messer et al., 1995; Sharp et al., 2006). Expected *a-posteriori* scoring was applied to extract factor scores, providing an estimate of the latent trait. The test information function (TIF), individual item information curves (IICs) and item characteristic curves (ICCs) assessed measurement precision (see supplementary material, page 2). Model fit was evaluated using Root-Mean-Square Error of Approximation (RMSEA), Comparative Fit Index (CFI) and Tucker-Lewis Index (TLI). Factor scores were then converted into T-scores, using the formula T = 50 + (Factor Score * 10) (de Beurs et al., 2022; McCall, 1922). Invariance analyses of SMFQ IRT-based T-scores between males and females were conducted to examine measurement equivalence across sexes (Table S1).

#### ROC curves

Receiver Operating Characteristic (ROC) analyses assessed the diagnostic performance of the SMFQ against DAWBA-based diagnoses, using both sum and IRT-based T-scores. Area under the curve (AUC) values were interpreted as low (≤0.7), moderate (0.7–0.9), or high (>0.9) (Henderson, 1993). Sensitivity and specificity were derived from ROC curves, and cut-offs were determined using the Youden Index (J = Sensitivity + Specificity - 1), ranging from 0 (no diagnostic ability) to 1 (perfect performance; Youden, 1950).

#### Diagnostic test properties

Post-test probabilities were calculated using Bayes’ theorem (Edwards, 1980; Fagan, 1975), incorporating weighted prevalence (pre-test probability) and ILRs for SMFQ T-score intervals. Unlike dichotomous likelihood ratios, ILRs are derived for test result ranges, minimizing information loss (Brown & Reeves, 2003; Gallagher, 1998). ILRs were computed for 5-point T-score intervals, or 10-point intervals at the extremes, ensuring sufficient cases in each group and merging similar intervals with overlapping 95% confidence intervals (Furukawa et al., 1997; Peirce & Cornell, 1993; Schmitz et al., 2000). Additional details are in supplementary material (page 2).

Logistic regression analyses evaluated whether the SMFQ T-score intervals improved prediction beyond the dichotomous operationalization obtained from ROC curves (Simel et al., 1993; Sugawara et al., 2017). A base model included only the dichotomous predictor. Subsequently, a second model was fitted adding T-score intervals. Models were compared using a chi-square test with 1 degree of freedom, under the null hypothesis that intervals did not enhance predictive accuracy. Additional logistic regression compared sum scores versus IRT-based T-scores (see supplementary material, page 2).

#### Sensitivity analysis

We estimated ROC curves using SMFQ IRT-based T-scores for all conditions excluding comorbidity with MDD. This was carried out to test whether the transdiagnostic association with other conditions, if any, could be due to MDD comorbidity.

Data analysis was conducted using RStudio version 4.2.1 (R Core Team, 2022) and MedCalc version 22.032 (MedCalc Statistical Software, 2024). The following R packages were utilized: *dplyr* for data manipulation (Wickham et al., 2023), *table1* for descriptive statistics (Rich, 2021), *mirt* for IRT analysis (Chalmers, 2012), *pROC* for ROC curves (Robin et al., 2011), *gtsummary* for summary tables (Sjoberg et al., 2021), *survey* for weighted prevalence (Lumley, 2004), *epiR* for epidemiological analysis (Stevenson et al., 2021), *stats* for analysis of variance (R Core Team, 2022), and *prevalence* to adjust prevalence for test properties (Devleesschauwer et al., 2022). MedCalc was utilized to generate ILRs and for ROC curves (MedCalc Statistical Software, 2024).

## Results

### Descriptive statistics

Of the 1905 participants at the 8-year follow-up, 1633 (85.7%) responded to the SMFQ, of whom 1582 (96.88%) completed all items. The sample was 54.4% male, with a mean age of 18.1 years (standard deviation [SD] = 1.98). An independent-samples t-test showed a significant sex difference in SMFQ scores. Females scored higher on the SMFQ (M = 6.85, SD = 6.9) than males (M = 4.35, SD = 5.27); t(1362.7) = 8.00, p < 0.001.

Any psychiatric condition was present in 28.2% of the sample (weighted prevalence [w] = 24.16%). MDD was the most common internalizing disorder (16.2%, w = 12.79%, n = 264), followed by PD/AG (6.5%, w = 5.68%), GAD (5.6%, w = 5.01%), SAD (3.1%, w = 2.76%), and PTSD (2.5%, w = 2.12%). ADHD and CD were present in 2.4% (w = 2.35%) and 2.9% (w = 2.71%), respectively. 71.8% had no psychiatric condition or missing data.

### Graded Response Model

#### Model Fit and Parameter Estimates

The GRM fit the data well (RMSEA = 0.047, 95% CI [0.041, 0.053], TLI = 0.989, CFI = 0.992), with an ω reliability of 0.944, supporting unidimensionality. Measurement invariance across sexes was accepted at both the metric and scalar levels (Table S1).

Discrimination parameters ranged from 0.904 to 4.174 (Table S2). Except for items 3, 4 and 7, all discrimination values were >2. Thresholds ranged from 0.162 to 1.045 for “Not true >= Sometimes” and from 1.293 to 2.393 for “Sometimes >= True”. Item 4 exhibited the lower threshold, indicating it is more readily endorsed by respondents. In contrast, item 2 demonstrated the higher threshold, suggesting it requires a higher level of the underlying trait for endorsement.

#### Test and Item Performance

The TIF indicated that most information was provided around 1.5 SD from the mean depressive symptom score (Figure S1). IICs (Figure S2) and ICCs (Figure S3) were also generated. Most items provided the most information for trait levels between −2.0 and 2.0 SD. Items 3 and 4 demonstrated shallow slopes, which indicates lower discriminating power for measuring depression.

#### IRT scores

IRT-derived scores strongly correlated with SMFQ sum scores (*r* = 0.94, p<0.001) (Figure S4). Sum scores ranged from 0 to 26 (mean = 5.51), while the IRT-based factor scores spanned from −1.11 to 2.76 (mean = 0.08). Factor scores were converted into T-scores, ranging from 39 to 78 (mean = 51.16). Chi-square model comparison revealed that adding IRT T-scores significantly improved predictions beyond sum scores for any condition, any internalizing condition, MDD, SAD, and PD/AG (Table S3) (Simel et al., 1993; Sugawara et al., 2017).

### Receiver Operating Characteristic (ROC) analyzes

Figure 1 presents ROC curves for SMFQ IRT-based T-scores. Sum score curves are available in the supplementary material (Figure S5). Prevalence estimates using IRT T-score cut-offs were adjusted to the population level using sensitivity and specificity parameters, resulting in values closely matching the weighted prevalence in the sample (Table S4). For IRT T-scores, sensitivity ranged from 51.06% (CD) to 84.31% (SAD), and specificity from 53.11% (ADHD) to 83.13% (GAD). The best cut-off for any diagnosis was >53 (sensitivity = 73.66%, 95% CI: 71.0 - 76.2; specificity = 75.22%, 95% CI: 71.0 - 79.1). For MDD, a cut-off point of >55 was identified (sensitivity = 76.52%, 95% CI: 70.9 - 81.5; specificity = 76.26%, 95% CI: 73.9 - 78.5). The highest AUC was observed when using the SMFQ to detect any internalizing condition (0.85, 95% CI: 0.83–0.86), and the lowest to detect ADHD (0.63, 95% CI: 0.61–0.66). For sum scores, the optimal cut-off to detect MDD was >6 (sensitivity = 73.73%, 95% CI: 67.9%–79.0%; specificity = 77.47%, 95% CI: 75.1%–79.7%) (Table S5). The traditional cut-off (≥8) showed lower sensitivity (64.71%, 95% CI: 58.5%–70.6%) but higher specificity (83.72%, 95% CI: 81.6%–85.7%).

**Figure 1.**
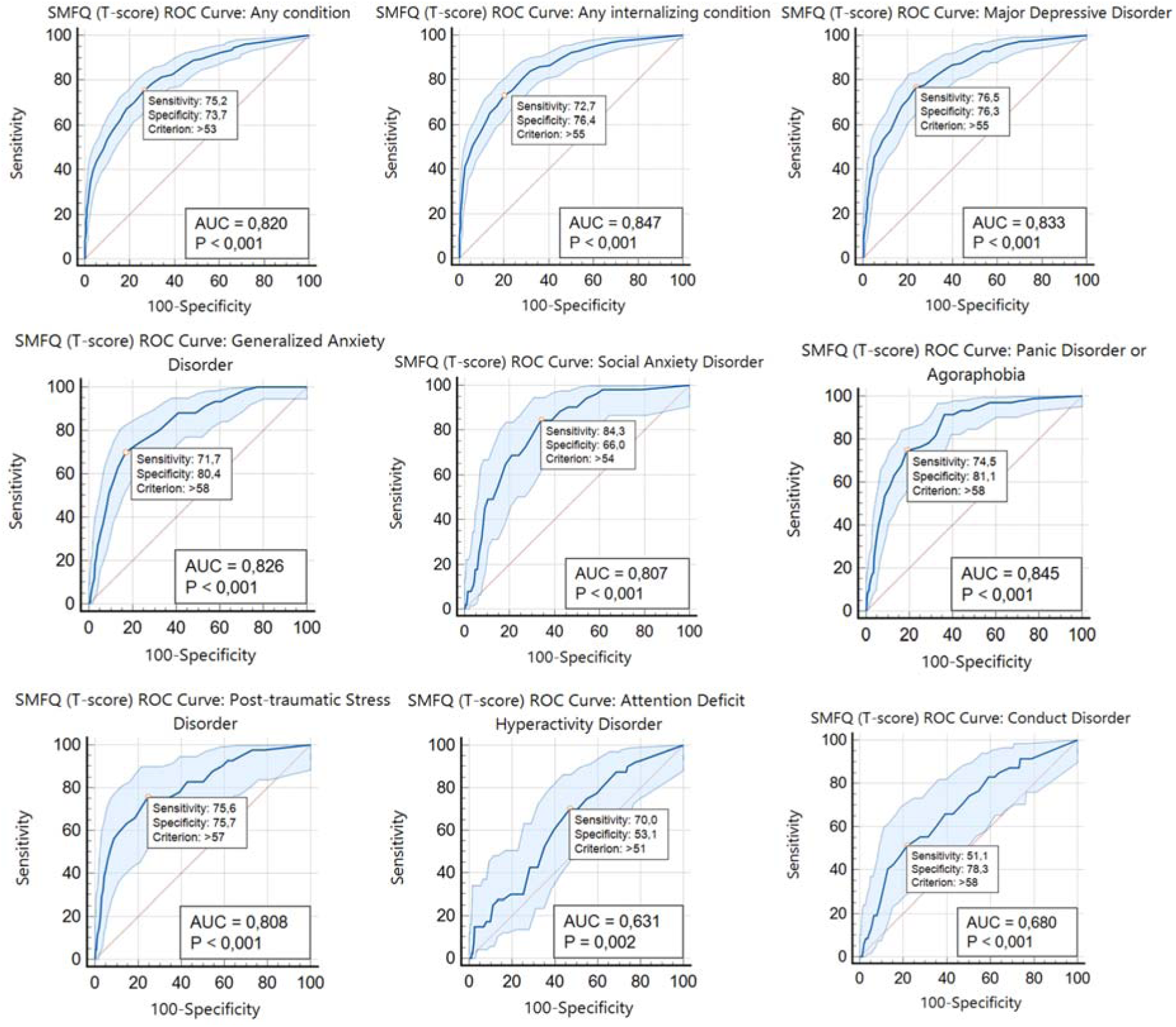
ROC Curves (T-scores) for all mental health conditions with prevalence ≥ 2% in this sample. **Note:** ROC curves were generated using the SMFQ T-score against DAWBA diagnosis. The highlighted point represents the chosen cut point for this sample and its sensitivity and specificity properties, respectively. SMFQ = Short Mood and Feelings Questionnaire. ROC = Receiver Operating Characteristic. AUC = Area Under the Curve.

### Diagnostic test properties for SMFQ interval cut-off scores

Table 1 presents ILRs and post-test probabilities for SMFQ score ranges. ADHD and CD are provided in the supplementary material (Table S6). For any condition (w = 24.16%), post-test probability surpassed pre-test probability for scores above the ROC-determined cut-off. A score >54 yielded a post-test probability of 35.72% (95% CI: 30.78% - 40.96%). This trend was also observed for any internalizing condition and PD/AG. For MDD, SAD, GAD, and PTSD, post-test probability decreased at higher levels. Trends for ADHD and CD could not be assessed due to the lack of cases at higher scores (Table S6). Based on these findings, we developed a web-based symptom calculator, available in American English (https://mheg.shinyapps.io/mfq-score-en/) and Brazilian Portuguese (https://mheg.shinyapps.io/mfq-score-main/). This tool enables clinicians to input SMFQ responses, obtain T-scores, and calculate post-test probabilities using ILRs. It also allows for the adjustment of pre-test parameters, facilitating its application across different contexts with comparable sample characteristics. This flexibility aims to enhance the external validity of the tool.

**Table 1.**
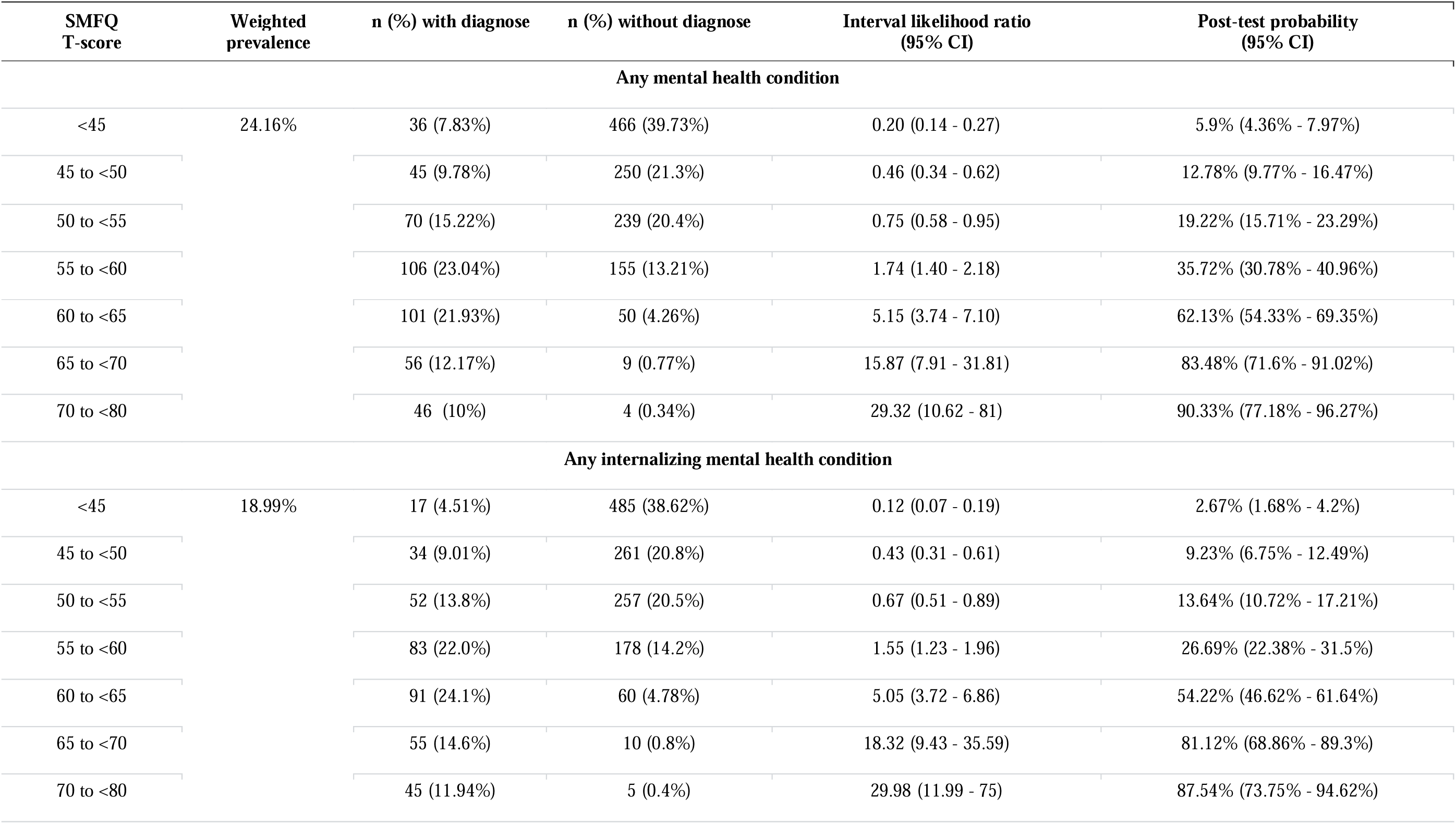
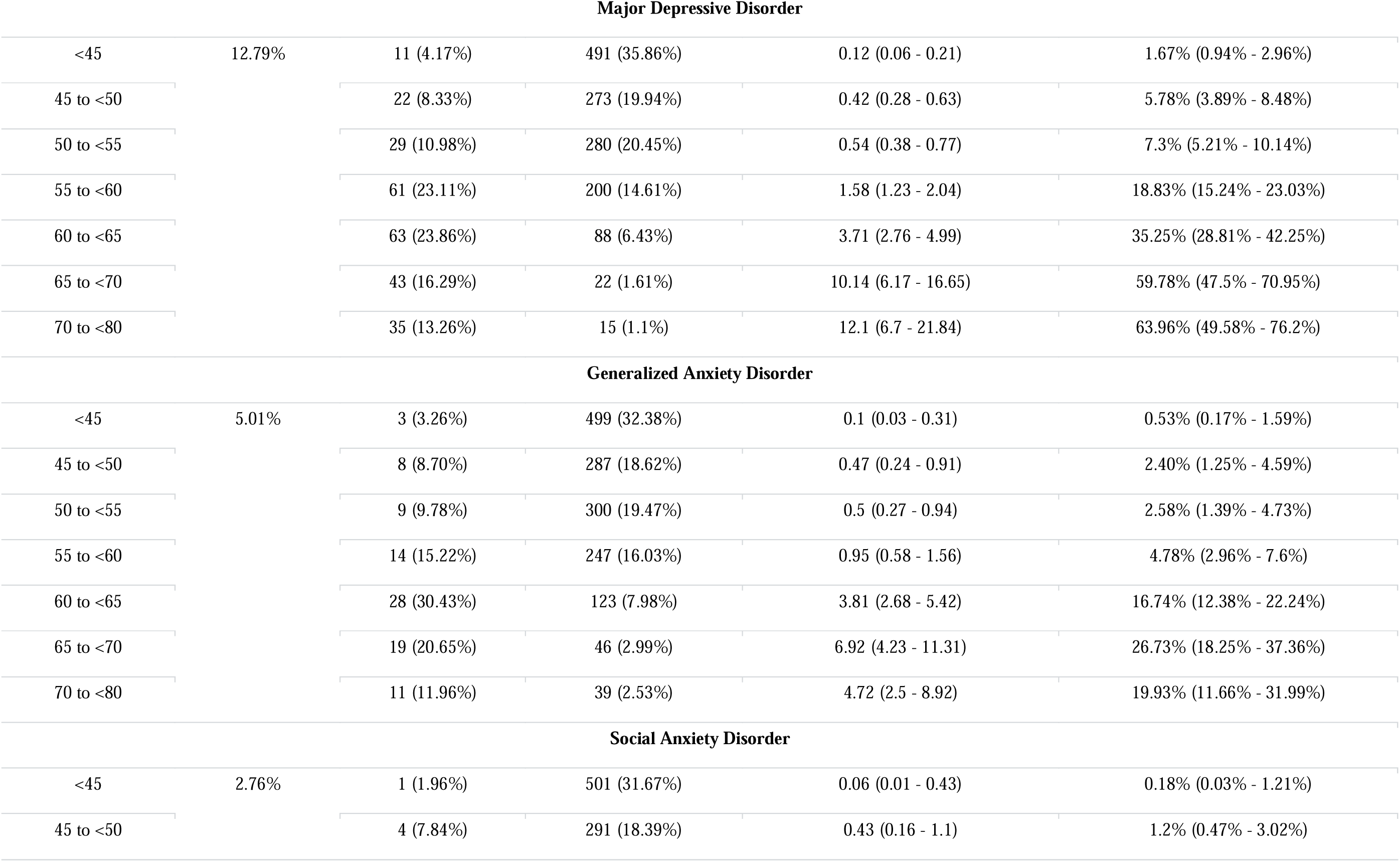
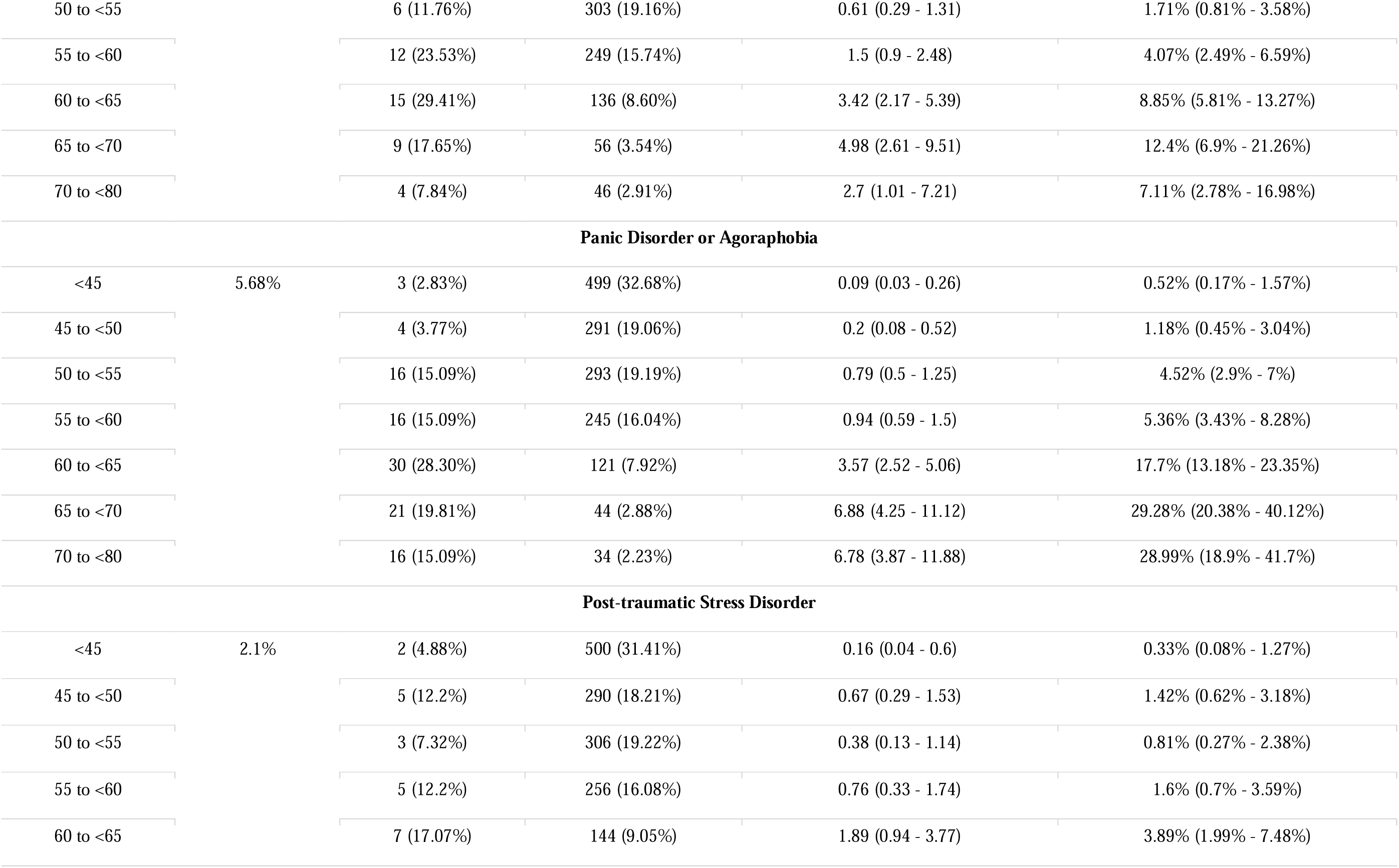
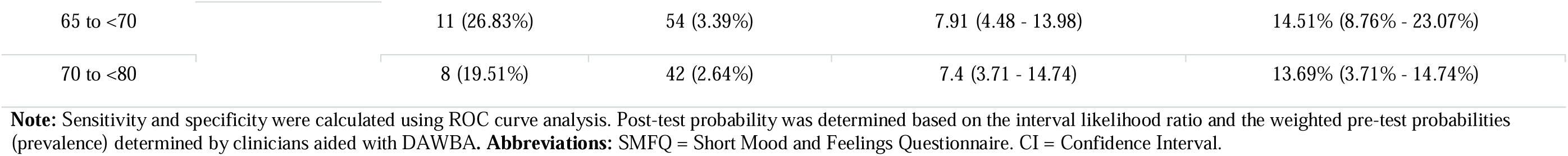
Post-test probabilities for various mental health conditions (n=1633).

The chi-square comparison indicated that interval predictors significantly improved prediction beyond using a single cut-off for any condition, any internalizing condition, MDD, GAD, SAD, PD/AG, and PTSD, but not for ADHD and CD (Table S7). These findings suggest that ILRs provide greater informational value than the dichotomous approach across most diagnostic categories.

### Sensitivity analysis

Excluding MDD comorbidity yielded similar ROC curves but with overall lower cut-off scores (Figure S6). For any internalizing condition, a >52 cut-off had 82.30% sensitivity (95% CI: 74.6%–88.8%) and 68.55% specificity (95% CI: 65.9%–71.1%) (Table S8). Including MDD reduced sensitivity (72.68%) but improved specificity (79.86%) (Table S4). AUC only slightly decreased from 0.85 to 0.82 (95% CI: 0.80 - 0.84) when excluding MDD, suggesting SMFQ retains its transdiagnostic utility.

## Discussion

In this study, we identified SMFQ cut-offs for MDD and other internalizing and externalizing conditions in a Brazilian urban school-based sample. We also evaluated the SMFQ’s ability to adjust pre-test probabilities using IRT-based T-score ranges and ILRs, instead of traditional dichotomization. The SMFQ exhibited high accuracy for detecting internalizing diagnoses, with higher T-scores corresponding to increased likelihood ratios and post-test probabilities. Lastly, to enhance clinical utility, we developed a calculator that estimates post-test probabilities based on SMFQ IRT-based T-scores.

We identified >6 as the optimal sum score cut-off for MDD, which is similar to the 8-point threshold proposed by Angold et al. (1995) for children but lower than that suggested by Turner et al. (2014) for our sample’s age range. Sum scores are widely used in clinical settings and generally perform well, assuming uniform item properties (Sijtsma et al., 2024). However, MDD symptoms (e.g., sad mood, suicidal ideation) represent distinct phenomena with varying risk and severity (Fried & Nesse, 2015). Our analysis showed SMFQ items differ in discrimination and threshold parameters, for instance.

IRT-based scores can refine clinical assessment by incorporating item discrimination and threshold parameters. Although sum and IRT-based scores are usually highly correlated, factor scores can vary by 1SD within the same 10-point score range (Damiano et al., 2023). At the traditional SMFQ threshold of 8 for MDD, factor scores ranged from 0.35 to 0.92, while at >6 (Table S5), they ranged from 0.28 to 0.75 (Figure S4). These findings underscore that identical sum scores may reflect substantial differences in factor scores. Our analysis suggests IRT improves detection of some conditions, including MDD (Table S3). To facilitate clinical implementation, we developed a calculator that converts SMFQ responses into standardized T-scores using IRT parameters. This standardization allows for harmonizing measurement scales and comparing disease severity in clinical and research settings (de Beurs et al., 2022). We also provided SMFQ cut-offs using this common metric to facilitate cross-scale comparisons.

Screening tools estimate an apparent prevalence, which differs from the true prevalence, established through gold-standard diagnostic criteria, due to the inherent uncertainty of such tools (Gonçalves Pacheco et al., 2023). Here, we adjusted the population prevalence estimates based on the sensitivity and specificity of the SMFQ. For instance, the apparent MDD prevalence from the SMFQ cut-off was 31.9%, but after adjustment, it aligned with the weighted prevalence (12.79%) at 15.2%. This highlights the need to account for diagnostic uncertainty, as failing to adjust test properties may distort prevalence estimates (Gonçalves Pacheco et al., 2023).

While dichotomous approaches like ROC curves are common in clinical assessments, they may oversimplify diagnostics. ILRs stratify scores into intervals, preserving symptom severity gradients and reducing information loss and misclassification (Brown & Reeves, 2003). We transformed T-scores into seven intervals, providing clinicians with actionable probabilities across the spectrum. Although ILRs remain underexplored in psychiatry, recent studies applied them to internalizing symptom scales (Blake et al., 2024; Sugawara et al., 2017). However, to our knowledge, no study has applied ILR to the MFQ or SMFQ. Our web-based tool enhances clinical decision-making by estimating post-test probabilities in real time, quickly converting raw scores into standardized T-scores based on latent estimates from a large sample. This enables personalized diagnosis and treatment based on probabilities rather than fixed categories. These innovations are especially valuable in LMICs, where resources are scarce, diagnostic uncertainty is high, and tailored care is crucial. Future research should assess whether ILR-based results influence clinical decisions, as choosing between ILRs and dichotomous LRs should prioritize clinical relevance over statistical concerns (Simel et al., 1993).

Our findings support transdiagnostic approaches (Fusar-Poli et al., 2019), particularly within the internalizing spectrum, as the SMFQ showed high accuracy in detecting these conditions. This spectrum, encompassing MDD, anxiety disorders, and related conditions, shares cognitive and emotional regulation deficits that contribute to their co-occurrence (Hankin et al., 2016). Other checklists, like the DSM Level 1 Cross-Cutting Symptom Measure, also exhibit transdiagnostic properties despite domain-specific intent (Pacheco et al., 2024). The SMFQ’s efficacy beyond its original purpose aligns with dimensional transdiagnostic models such as the Hierarchical Taxonomy of Psychopathology (HiTOP) (Kotov et al., 2021). Given the high comorbidity between internalizing and externalizing conditions (McGrath et al., 2020; Caspi et al., 2020), we tested whether the SMFQ could detect ADHD and CD, which was not confirmed. Still, its transdiagnostic utility within internalizing conditions is valuable, potentially streamlining diagnostics, reducing reliance on multiple scales, and aiding early intervention. Whether through dichotomous or interval-based methods, the SMFQ remained transdiagnostic within internalizing conditions.

The study must be understood in light of its limitations. First, the sample comprises young people, and results might not apply to children or older adults. Nonetheless, scores were calculated using weights to adjust for follow-up missingness and the BHRCS oversampling procedure, reducing sample bias. Second, small sample sizes for certain diagnostic intervals may have affected the accuracy of the ILRs, particularly for CD or ADHD. Lastly, the study relies on cross-sectional data, causal inferences and the assessment of SMFQ score stability over time. Future research should address these gaps by incorporating longitudinal data to assess the consistency of these findings over time and expanding the sample to better evaluate its applicability to a broader spectrum of psychopathology.

## Conclusion

This study estimated SMFQ cut-offs for Brazilian youth and demonstrated that ILRs enhance its ability to detect MDD. By applying IRT to generate T-scores and ILRs, we improved the SMFQ’s precision and flexibility beyond simple dichotomization. A web-based calculator further supports integrating these methods into clinical practice, enabling real-time, data-driven decisions. Our findings also suggest the SMFQ’s potential as a transdiagnostic screening tool for internalizing conditions, particularly in low-resource settings. Future research should examine the broader applicability of these methods across different populations, evaluate their ability to predict functional outcomes, and investigate the transdiagnostic potential of similar tools.

## Data Availability

Data dictionary is available at https://osf.io/ktz5h/wiki/Data%20Dictionaries/ and https://osf.io/w3jr4 to direct download. Individual-level data is available upon request to the Brazilian High-Risk Cohort Study research committee, by following the instructions and filling the research form available at https://osf.io/ktz5h/wiki/home/.

## Acknowledgements

Gabriele dos Santos Jobim was supported by FIPE/UFSM 2024 scholarship and CNPq/UFSM 2024 scholarship, granted to prof. Mauricio Scopel Hoffmann.

Dr. João V. do Amaral was supported by the Coordenação de Aperfeiçoamento de Pessoal de Nível Superior – Brasil (CAPES) – Finance Code 001.

Dr. Mauricio Scopel Hoffmann was supported by the United States National Institutes of Health grant R01MH120482 under his post-doctoral fellowship at UFRGS.

## CRediT authorship contribution statement

GSJ: Conceptualization, study design, data curation, formal analysis, investigation, methodology, project administration, software, validation, visualisation, writing – original draft, and writing – review & editing. JVA: Conceptualization, formal analysis, methodology, software, writing – review & editing. JPGP: Conceptualization, study design, methodology, writing – review & editing. AG: Data curation, funding acquisition, writing – review & editing. ECM: Data curation, funding acquisition, writing – review & editing. RAB: Funding acquisition, writing – review & editing. PMP: Data curation, funding acquisition, writing – review & editing. LAR: Funding acquisition, writing – review & editing. GAS: Data curation, funding acquisition, methodology, and writing – review & editing. MSH: Conceptualization, methodology, formal analysis, investigation, data curation, writing – original draft, writing – review & editing, and supervision.

JVA and MSH directly accessed and verified the underlying data reported in the manuscript.

All authors read and approved the final version of the manuscript and had full access to all the data in the study and had final responsibility for the decision to submit for publication.

## Data sharing statement

Data dictionary is available at https://osf.io/ktz5h/wiki/Data%20Dictionaries/ and https://osf.io/w3jr4 to direct download. Individual-level data is available upon request to the Brazilian High-Risk Cohort Study research committee, by following the instructions and filling the research form available at https://osf.io/ktz5h/wiki/home/. Study design and ethical details can be found elsewhere (Salum et al., 2015).

