## Supplementary material for "Clinical properties of the Short Mood and Feelings Questionnaire: Development of a free calculator based on a Brazilian High-Risk Cohort Study"

Content:

| Statistical analysis | 2 |
| --- | --- |
| Table S1 - Measurement invariance of SMFQ across sex groups. | 3 |
| Table S2. SMFQ item response theory (graded response model) parameters | 4 |
| Table S3 - Logistic regression models using sum scores and IRT-based T-scores. | 5 |
| Table S4 - Sensitivity, specificity, predictive values, likelihood ratios, and accuracy for each cut-off point of the SMFQ (IRT-based T-score), by diagnoses | 6 |
| Table S5 - Sensitivity, specificity, predictive values, likelihood ratios, and accuracy for each diagnosis, assessed by SMFQ sum scores. | 8 |
| Table S6 - Diagnostic test properties for ADHD and CD (n=1633). | 9 |
| Table S7 - Logistic regression models using dichotomous and interval predictors | 10 |
| Table S8 - Sensitivity, specificity, predictive values, likelihood ratios, and accuracy for each cut-off point of the SMFQ (T-score), by each diagnosis, excluding major depressive disorder diagnosis. | 11 |
| Figure S1 - Test information curve (solid line) and standard error (dashed line) of the Short Mood and Feelings Questionnaire (SMFQ). | 12 |
| Figure S2 - Item information curves for the 13 items of the Short Mood and Feelings Questionnaire (SMFQ). | 12 |
| Figure S3 - Item characteristic curves for the 13 items of the Short Mood and Feelings Questionnaire (SMFQ). | 13 |
| Figure S4 - Correlation between Short Mood and Feelings Questionnaire (SMFQ) total and IRT-based factor score (T-score). | 13 |
| Figure S5 - SMFQ (sum scores) ROC Curves for all mental health conditions with prevalence ≥ 2% in this sample. | 14 |
| Figure S6 - SMFQ (T-score) ROC Curves for all mental health conditions with prevalence ≥ 2% in this sample, excluding major depressive disorder diagnosis. | 15 |

**Statistical analysis**

Item response theory (IRT) analyses employed a graded response model (GRM) designed for categorical data with ordered responses. The GRM uses two key parameters: discrimination and thresholds. Discrimination measures how well an item differentiates between individuals with different levels of the underlying trait; higher values indicate greater sensitivity. Threshold parameters represent the points on the latent trait continuum where a respondent has a 50% probability of endorsing the next higher response category. For an item with k response categories, there are "k-1" thresholds (b1, b2, ..., b_(k-1)_). Sampling and attrition weights were applied in the analysis to ensure representativeness of the population. The TIF quantifies the overall amount of information provided by the test at various levels of the latent trait, with peaks indicating high precision and dips at endpoints showing less reliable measurement. Individual item information curves (IICs) show where each item contributes most to measurement precision. Item characteristic curves (ICCs) visually represent the probability of endorsing an item across different levels of the latent trait. Shallower slopes in ICCs indicate lower item discrimination. In terms of model fit, we considered a RMSEA equal or below 0.06 and CFI and TLI above 0.95 as indicators of a good fit (Brown, 2015).

To calculate ILRs, IRT T-scores were grouped into 5-point intervals, or 10-point intervals at the extremes, based on the recommendation to ensure a sufficient number of cases and noncases in each group so that the ILR maintains a monotonic relationship. It is also advised to combine intervals where the ILRs are similar and their 95% confidence intervals significantly overlap (Furukawa et al., 1997; Peirce & Cornell, 1993; Schmitz et al., 2000). For each interval, the likelihood ratio is calculated as the probability of a given test result interval in those with the disease divided by the probability of the same result interval in those without the disease (Brown & Reeves, 2003). Specifically, the ILR for an interval [a, b] is defined as the probability of the test result being within the interval [a, b] given the presence of the disease, divided by the probability of the test result being within the same interval given the absence of the disease. These ILRs were then used to generate post-test probabilities, applying Bayes' theorem (Edwards, 1980; Fagan, 1975). This method transforms the pre-test probability into pre-test odds, which are then adjusted using ILRs for each SMFQ T-score 5-point interval (35 to <40 to 75 to <80) to obtain post-test odds. Finally, these post-test odds are converted back into post-test probabilities. This procedure was done for all mental health conditions included in the study.

Logistic regression analyses were conducted to compare the predictive performance of sum scores versus IRT-based T-scores of the SMFQ. The base model included only the sum score predictor, and the second model incorporated both the sum score and IRT-based T-score predictors. The difference between the two models was evaluated using a chi-square test with 1 degree of freedom, under the null hypothesis that the inclusion of IRT-derived T-scores did not enhance predictive accuracy. A non-significant result would suggest that the inclusion of IRT did not contribute additional informative value beyond what is captured by sum scores.

| **Table S1.** Measurement invariance of SMFQ across sex groups | | | | | | | | | | |
| --- | --- | --- | --- | --- | --- | --- | --- | --- | --- | --- |
| Sample in each group | Invariance | M2 | *df* | RMSEA | CFI | Model comparison | ΔM2 (Δdf) | Δ CFI | Δ RMSEA | Decision |
| Female = 735  Male = 847 | Configural | 386.969 | 130 | 0.035 | 0.992 |  |  |  |  |  |
|  | Metric | 393.161 | 143 | 0.033 | 0.992 | Configural | 6.192 (13) | 0.000 | 0.002 | Accept |
|  | Scalar | 691.386 | 169 | 0.044 | 0.983 | Metric | 298.225 (26) | 0.009 | 0.011 | Accept |
| **Note:** Invariance decision is based on ΔCFI < 0.010 supplemented by ΔRMSEA < 0.015 or ΔSRMR < 0.010. M2, M2 model fit statistic; DF, degrees of freedom; RMSEA, Root Mean Square Error of Approximation; CFI, Comparative Fit Index; TLI, Tucker-Lewis Index; SRMR, Standardized Root Mean-square Residual; Δ, differences between fit index. **, p < 0.01; ***, p < 0.001 | | | | | | | | | | |

| **Table S2.** SMFQ item response theory (graded response model) parameters | | | |
| --- | --- | --- | --- |
|  |  | **b (thresholds)** | |
| **SMFQ item** | **a (discrimination)** | **Not true >= sometimes** | **Sometimes >= True** |
| 1. I felt miserable or unhappy | 2.648 | 0.347 | 1.524 |
| 2. I didn’t enjoy anything at all | 2.015 | 1.045 | 2.298 |
| 3. I felt so tired I just sat around and did nothing | 1.726 | 0.302 | 1.555 |
| 4. I was very restless | 0.904 | 0.162 | 2.393 |
| 5. I felt I was no good any more | 4.057 | 0.882 | 1.605 |
| 6. I cried a lot | 2.621 | 0.628 | 1.439 |
| 7. I found it hard to think properly or concentrate | 1.819 | 0.259 | 1.717 |
| 8. I hated myself | 4.174 | 0.935 | 1.639 |
| 9. I was a bad person | 3.858 | 0.941 | 1.739 |
| 10. I felt lonely | 4.023 | 0.423 | 1.293 |
| 11. I thought nobody really loved me | 3.406 | 0.895 | 1.655 |
| 12. I thought I could never be as good as other kids | 2.733 | 0.547 | 1.589 |
| 13. I did everything wrong | 3.177 | 0.835 | 1.718 |

**Note:** The self-report version of the Short Mood and Feelings Questionnaire (SMFQ) comprises 13 items (Angold & Costello, 1987). Copyright Adrian Angold & Elizabeth J. Costello, 1987; Developed Epidemiology Program, Duke University. Reproduced with permission from developer, may be reproduced for use with one’s own patients

| **Table S3**. Logistic regression models using sum scores and IRT-based T-scores. | | | | | | | |
| --- | --- | --- | --- | --- | --- | --- | --- |
|  | Logistic regressions (sum score predictor only) | | Logistic regressions  (sum score + IRT T-score predictor) | | | | Chi-square model comparison (p-value) |
|  |  |  | Sum score predictor | | IRT T-score predictor | |  |
| Diagnosis | OR | p | OR | p | OR | p |  |
| Any condition | 1.25 | **<0.001** | 1.14 | **<0.001** | 1.07 | **0.008** | **0.007** |
| Any internalizing condition | 1.26 | **<0.001** | 1.11 | **0.007** | 1.11 | **<0.001** | **<0.001** |
| Major Depressive Disorder | 1.21 | **<0.001** | 1.07 | 0.118 | 1.11 | **0.001** | **<0.001** |
| Generalized Anxiety Disorder | 1.17 | **<0.001** | 1.10 | 0.105 | 1.05 | 0.297 | 0.289 |
| Social Anxiety Disorder | 1.14 | **<0.001** | 0.95 | 0.510 | 1.17 | **0.024** | **0.016** |
| Panic Disorder or Agoraphobia | 1.18 | **<0.001** | 1.03 | 0.616 | 1.13 | **0.020** | **0.015** |
| Post-traumatic Stress Disorder | 1.17 | **<0.001** | 1.17 | 0.072 | 1.00 | 0.980 | 0.98 |
| Attention Deficit Hyperactivity Disorder | 1.06 | **0.005** | 1.00 | 0.981 | 1.05 | 0.422 | 0.418 |
| Conduct Disorder | 1.09 | **<0.001** | 1.06 | 0.372 | 1.02 | 0.752 | 0.751 |

**Note:** The sum score predictor represents the sum of all 13 SMFQ items, while the IRT score predictor is the conversion of IRT factor scores into T-scores. These predictors were compared in logistic regression models to determine which more accurately predicted the presence of a diagnosis. The difference between these models was tested using a chi-square test with 1 degree of freedom, under the null hypothesis that the inclusion of IRT T-scores did not improve predictive accuracy beyond using sum scores. **Abbreviations:** OR=Odds Ratio.

| **Table S4.** Sensitivity, specificity, predictive values, likelihood ratios, and accuracy for each cut-off point of the SMFQ (IRT-based T-score), by diagnoses | | | | | | | | | | | | | | | | |
| --- | --- | --- | --- | --- | --- | --- | --- | --- | --- | --- | --- | --- | --- | --- | --- | --- |
| **Diagnosis (weighted prevalence)** | **Cut-off** | **Prevalence; adjusted prevalence (95% CI)*** | **Sensitivity**  **(95% CI)** | | **Specificity**  **(95% CI)** | | **Youden Index (95% CI)** | **Positive predictive value (95% CI)** | | **Negative predictive value (95% CI)** | | **Positive likelihood ratio (95% CI)** | | **Negative likelihood ratio (95% CI)** | | **AUC**  **(95% CI)** |
| Any condition  (24.16%) | >53 | 39.62%; 27.8% (23.1% - 32.7%) | 75.22%  (71% - 79.1%) | | 73.66%  (71% - 76.2%) | | 48.87%  (0.44% - 0.53%) | 47.6  (44.9 - 50.4) | | 90.3  (88.8 - 91.7) | | 2.86  (2.56 - 3.18) | | 0.34  (0.29 - 0.40) | | 0.82  (0.8 - 0.84) |
| Any internalizing condition  (18.99%) | >55 | 31.9%; 22.5% (18.2% - 26.8%) | 72.68%  (67.9% - 77.1%) | | 79.86%  (77.5% - 82%) | | 52.54%  (0.47% - 0.56%) | 45.8  (42.7 - 49.0) | | 92.6  (91.3 - 93.6) | | 3.61  (3.18 - 4.09) | | 0.34  (0.29 - 0.40) | | 0.85  (0.83 - 0.86) |
| Major Depressive Disorder  (12.79%) | >55 | 31.9%; 15.2% (10.9% - 19.5%) | 76.52%  (70.9% - 81.5%) | | 76.26%  (73.9% - 78.5%) | | 52.78%  (47.48% - 57.97%) | 32.1  (29.6 - 34.7) | | 95.7  (94.7 - 96.5) | | 3.22  (2.87 - 3.62) | | 0.31  (0.25 - 0.38) | | 0.83  (0.81 - 0.85) |
| Generalized Anxiety Disorder  (5.01%) | >59 | 19.66%; 5.1%  (1.6% - 8.8%) | 69.57%  (59.1% - 78.7%) | | 83.13%  (81.2% - 85%) | | 52.69%  (0.42% - 0.60%) | 17.9  (15.4 - 20.6) | | 98.1  (97.4 - 98.6) | | 4.12  (3.46 - 4.91) | | 0.37  (0.27 - 0.50) | | 0.83  (0.81 - 0.84) |
| Social Anxiety Disorder  (2.76%) | >54 | 35.33%; 3.3%  (0.2% - 7.5%) | 84.31%  (71.4% - 93%) | | 65.99%  (63.6% - 68.3%) | | 50.31%  (0.36% - 0.58%) | 6.6  (5.8 - 7.5) | | 99.3  (98.7 - 99.6) | | 2.48  (2.16 - 2.84) | | 0.24  (0.13 - 0.45) | | 0.81  (0.79 - 0.83) |
| Panic Disorder or Agoraphobia  (5.68%) | >58 | 22.35%; 6.2%  (2.5% - 9.9%) | 74.53%  (65.1% - 82.5%) | | 81.07%  (79% - 83%) | | 55.6%  (0.47% - 0.61%) | 19.2  (16.9 - 21.6) | | 98.1  (97.4 - 98.7) | | 3.94  (3.38 - 4.59) | | 0.31  (0.23 - 0.44) | | 0.84  (0.83 - 0.86) |
| Post-traumatic Stress Disorder (2.12%) | >57 | 25.29%; 2.9%  (0.2% - 6.7%) | 75.61%  (59.7% - 87.6%) | | 75.69%  (73.5% - 77.8%) | | 51.30%  (0.36% - 0.62%) | 6.3  (5.2 - 7.5) | | 99.3  (98.8 - 99.6) | | 3.11  (2.56 - 3.78) | | 0.32  (0.19 - 0.55) | | 0.81  (0.79 - 0.83) |
| Attention Deficit Hyperactivity Disorder  (2.35%) | >51 | 46.85%; 4%  (0.2% - 11.5%) | 70%  (53.5% - 83.4%) | | 53.11%  (50.6% - 55.6%) | | 23.11%  (0.1% - 0.32%) | 3.5  (2.8 - 4.2) | | 98.7  (97.9 - 99.2) | | 1.49  (1.21 - 1.84) | | 0.56  (0.35 - 0.91) | | 0.63  (0.61 to 0.66) |
| Conduct Disorder  (2.71%) | >58 | 22.35%; 3.4%  (0.1% - 9.1%) | 51.06%  (36.1% - 65.9%) | | 78.31%  (76.2% - 80.3%) | | 29.37%  (0.15% - 0.4%) | 6.2  (4.7 - 8.1) | | 98.3  (97.7 - 98.7) | | 2.35  (1.75 - 3.16) | | 0.62  (0.47 - 0.84) | | 0.68  (0.66 to 0.7) |

**Note:** Optimal cut-off points were determined by the analysis of ROC (Receiver Operating Characteristic) curves (i.e., Youden index) for the SMFQ T-score against DAWBA diagnosis, balancing sensitivity and specificity. *Prevalence was calculated using each recommended cut-off and then adjusted for the population level using sensitivity and specificity parameters. **Abbreviations:** CI=Confidence Interval; AUC=Area under the curve.

| **Table S5.** Sensitivity, specificity, predictive values, likelihood ratios, and accuracy for each diagnosis, assessed by SMFQ sum scores. | | | | | | | | | | | | | | | |
| --- | --- | --- | --- | --- | --- | --- | --- | --- | --- | --- | --- | --- | --- | --- | --- |
| **Diagnosis** | **Cut-off** | **Sensitivity**  **(95% CI)** | | **Specificity**  **(95% CI)** | | **Youden Index (95% CI)** | **Positive predictive value (95% CI)** | | **Negative predictive value (95% CI)** | | **Positive likelihood ratio (95% CI)** | | **Negative likelihood ratio (95% CI)** | | **AUC**  **(95% CI)** |
| Any disorder | >4 | 77.03%  (72.8% - 80.9% ) | | 73.9%  (71.2% - 76.4%) | | 50.93%  (46.31% - 55.56%) | 48.5  (45.7 - 51.2) | | 91.0  (89.5 - 92.3) | | 2.95  (2.64 - 3.30) | | 0.31  (0.26 - 0.37) | | 0.82  (0.8 - 0.84) |
| Any internalizing disorder | >4 | 81.69%  (77.3% - 85.5%) | | 72.04%  (69.4% - 74.5%) | | 53.73%  (48.87% - 58.36%) | 40.6  (38.2 - 43.1) | | 94.4  (93.1 - 95.4) | | 2.92  (2.64 - 3.24) | | 0.25  (0.20 - 0.32) | | 0.85  (0.83 - 0.86) |
| Major Depressive Disorder | >6 | 73.73%  (67.9% - 79.0%) | | 77.47%  (75.1% - 79.7%) | | 51.19%  (44.87% - 55.67%) | 32.4  (29.8 - 35.2) | | 95.3  (94.2 - 96.1) | | 3.27  (2.89 - 3.70) | | 0.34  (0.28 - 0.42) | | 0.83  (0.81 - 0.85) |
| Generalized Anxiety Disorder | >9 | 72.53%  (62.2% - 81.4%) | | 81.62%  (79.6% - 83.6%) | | 54.15%  (43.77% - 62.61%) | 17.2  (15 - 19.7) | | 98.3  (97.6 - 98.7) | | 3.95  (3.34 - 4.66) | | 0.34  (0.24 - 0.47) | | 0.83  (0.81 - 0.85) |
| Social Anxiety Disorder | >4 | 86%  (73.3% - 94.2%) | | 61.1%  (58.6% - 63.5%) | | 47.10%  (34.3% to 54.25%) | 5.9  (5.2 - 6.7) | | 99.4  (98.7 - 99.7) | | 2.21  (1.94 - 2.51) | | 0.23  (0.12 - 0.46) | | 0.8  (0.78 - 0.82) |
| Panic Disorder or Agoraphobia | >7 | 77.14  (67.9% - 84.8%) | | 76.44%  (74.2% - 78.6%) | | 53.58%  (45.05% - 59.64%) | 16.5  (14.6 - 18.5) | | 98.2  (97.5 - 98.8) | | 3.27  (2.85 - 3.76) | | 0.3  (0.21 - 0.43) | | 0.84  (0.82 - 0.86) |
| Post-traumatic Stress Disorder | >10 | 68.29%  (51.9% - 81.9%) | | 82.09%  (80.1% - 84%) | | 50.38%  (33.59% - 62.36%) | 7.6  (6.1 - 9.4) | | 99.2  (98.7 - 99.5) | | 3.81  (3.02 - 4.82) | | 0.39  (0.25 - 0.61) | | 0.81  (0.79 - 0.83) |
| Attention Deficit Hyperactivity Disorder | >3 | 71.05%  (54.1% - 84.6%) | | 52.85%  (50.3% - 55.4%) | | 23.9%  (10.04% - 35.75%) | 3.5  (2.9 - 4.3) | | 98.7  (97.9 - 99.2) | | 1.51  (1.22 - 1.86) | | 0.55  (0.33 - 0.9) | | 0.63  (0.61 - 0.66) |
| Conduct Disorder | >11 | 45.65%  (30.9% - 61%) | | 84.31%  (82.4% - 86.1%) | | 29.96%  (16.33% - 41.08%) | 7.5  (5.5 - 10.2) | | 98.2  (97.7 - 98.6) | | 2.91  (2.08 - 4.07) | | 0.64  (0.49 - 0.84) | | 0.68  (0.65 - 0.7) |

**Note:** Optimal cut points were determined by the analysis of ROC (Receiver Operating Characteristic) curves, balancing sensitivity and specificity. **Abbreviations:** CI=Confidence Interval; AUC=Area under the curve.

| **Table S6.** Diagnostic test properties for ADHD and CD (n=1633). | | | | | |
| --- | --- | --- | --- | --- | --- |
| **Cut-off** | **Weighted prevalence** | **n (%) with diagnose** | **n (%) without diagnose** | **Likelihood ratio**  **(95% CI)** | **Post-test probability**  **(95% CI)** |
| **Attention Deficit Hyperactivity Disorder** | | | | | |
| <45 | 2.35% | 5 (12.5%) | 497 (31.2%) | 0.4 (0.18 - 0.91) | 0.96% (0.42% - 2.15%) |
| 45 to <50 |  | 7 (17.5%) | 288 (18.08%) | 0.97 (0.49 - 1.91) | 2.28% (1.16% - 4.4%) |
| 50 to <55 |  | 11 (27.5%) | 298 (18.71%) | 1.47 (0.88 - 2.46) | 3.42% (2.07% - 5.58%) |
| 55 to <60 |  | 6 (15%) | 255 (16.01%) | 0.94 (0.44 - 1.98) | 2.2% (1.06% - 4.54%) |
| 60 to <65 |  | 5 (12.5%) | 146 (9.17%) | 1.36 (0.59 - 3.14) | 3.18% (1.4% - 7.03%) |
| 65 to <70 |  | 0 (0.00%) | 65 (4.08%) | 0 (0 - 4.86) | 0 (0 - 10.47%) |
| 70 to <80 |  | 6 (15%) | 44 (2.76%) | 5.43 (2.46 - 12) | 11.56% (5.58% - 22.41%) |
| **Conduct Disorder** | | | | | |
| <45 | 2.71% | 6 (12.77%) | 496 (31.27%) | 0.41 (0.19 - 0.86) | 1.12% (0.54% - 2.35%) |
| 45 to <50 |  | 6 (12.77%) | 289 (18.22%) | 0.7 (0.33 - 1.49) | 1.92% (0.91% - 3.98%) |
| 50 to <55 |  | 9 (19.15%) | 300 (18.91%) | 1.01 (0.56 - 1.84) | 2.74% (1.53% - 4.87%) |
| 55 to <60 |  | 6 (12.77%) | 255 (16.08%) | 0.79 (0.37 - 1.69) | 2.16% (1.03% - 4.5%) |
| 60 to <65 |  | 11 (23.40%) | 140 (8.83%) | 2.65 (1.54 - 4.55) | 6.88% (4.12% - 11.26%) |
| 65 to <70 |  | 5 (10.64%) | 60 (3.78%) | 2.81 (1.18 - 6.68) | 7.26% (3.19% - 15.69%) |
| 70 to <80 |  | 4 (8.51%) | 46 (2.9%) | 2.93 (1.1 - 7.82) | 7.56% (2.98% - 17.88%) |

**Note:** Sensitivity and specificity were calculated using ROC curve analysis. Post-test probability was determined based on the interval likelihood ratio and the weighted pre-test probabilities (prevalence) determined by clinicians aided with DAWBA**. Abbreviations:** CI = Confidence Interval; SMFQ = Short Mood and Feelings Questionnaire.

| **Table S7.** Logistic regression models using dichotomous and interval predictors | | | | | | | |
| --- | --- | --- | --- | --- | --- | --- | --- |
|  | Logistic regressions (dichotomous predictor only) | | Logistic regressions  (dichotomous + interval predictor) | | | | Chi-square model comparison (p-value) |
|  |  |  | Dichotomous predictor | | Interval predictor | |  |
| Diagnosis | OR | p | OR | p | OR | p |  |
| Any condition | 8.51 | **<0.001** | 1.16 | 0.493 | 2.11 | **<0.001** | **<0.001** |
| Any internalizing condition | 10.48 | **<0.001** | 1.01 | 0.965 | 2.42 | **<0.001** | **<0.001** |
| Major Depressive Disorder | 10.20 | **<0.001** | 1.49 | 0.124 | 1.99 | **<0.001** | **<0.001** |
| Generalized Anxiety Disorder | 11.42 | **<0.001** | 2.6 | **0.025** | 1.61 | **<0.001** | **<0.001** |
| Social Anxiety Disorder | 10.55 | **<0.001** | 2.9 | 0.066 | 1.52 | **0.002** | **0.003** |
| Panic Disorder or Agoraphobia | 12.70 | **<0.001** | 3.07 | **0.004** | 1.60 | **<0.001** | **<0.001** |
| Post-traumatic Stress Disorder | 9.82 | **<0.001** | 1.62 | 0.417 | 1.83 | **<0.001** | **<0.001** |
| Attention Deficit Hyperactivity Disorder | 2.71 | **0.004** | 1.78 | 0.296 | 1.16 | 0.310 | 0.316 |
| Conduct Disorder | 3.81 | **<0.001** | 2.68 | 0.071 | 1.12 | 0.442 | 0.444 |

**Note:** The dichotomous predictor was based on SMFQ cut-offs identified through ROC analysis for each condition, while the interval predictor was derived from IRT-based SMFQ T-scores using interval likelihood ratio analysis. These predictors were compared in logistic regression models to determine which more accurately predicted the presence of a diagnosis. The difference between these models was tested using a chi-square test with 1 degree of freedom, under the null hypothesis that the inclusion of intervals did not improve predictive accuracy beyond the single cut-off point. **Abbreviations:** OR=Odds Ratio.

| **Table S8.** Sensitivity, specificity, predictive values, likelihood ratios, and accuracy for each cut-off point of the SMFQ (T-score), by each diagnosis, excluding major depressive disorder diagnosis. | | | | | | | | | | | | | | | |
| --- | --- | --- | --- | --- | --- | --- | --- | --- | --- | --- | --- | --- | --- | --- | --- |
| **Diagnosis** | **Cut-off** | **Sensitivity**  **(95% CI)** | | **Specificity**  **(95% CI)** | | **Youden Index (95% CI)** | **Positive predictive value (95% CI)** | | **Negative predictive value (95% CI)** | | **Positive likelihood ratio (95% CI)** | | **Negative likelihood ratio (95% CI)** | | **AUC (95% CI)** |
| Any disorder | >52 | 70.41%  (63.5% - 76.7%) | | 70.16%  (67.5% - 72.8%) | | 0.41%  (0.32% - 0.47%) | 42.9  (39.9 - 46.0) | | 88.2  (85.7 - 90.3) | | 2.36  (2.08 - 2.68) | | 0.42  (0.34 - 0.53) | | 0.76  (0.74 - 0.78) |
| Any internalizing disorder | >52 | 82.30%  (74% - 88.8%) | | 68.55%  (65.9% - 71.1%) | | 0.51%  (0.42% - 0.58%) | 38.0  (35.3 - 40.8) | | 94.3  (91.7 - 96.1) | | 2.62  (2.33 - 2.95) | | 0.26  (0.17 - 0.38) | | 0.82  (0.8 - 0.84) |
| Generalized Anxiety Disorder | >52 | 89.36%  (76.9% - 96.5%) | | 66.26%  (63.6% - 68.8%) | | 0.56%  (0.43% - 0.62%) | 12.3  (11.0 - 13.7) | | 99.2  (98.1 - 99.6) | | 2.65  (2.34 - 3) | | 0.16  (0.07 - 0.37) | | 0.85  (0.83 - 0.87) |
| Social Anxiety Disorder | >54 | 75.86%  (56.5% - 89.7%) | | 73.88%  (71.4% - 76.2%) | | 0.5%  (0.36% - 0.61%) | 7.6  (6.2 - 9.4) | | 99.1  (98.3 - 99.5) | | 2.9  (2.32 - 3.63) | | 0.33  (0.17 - 0.62) | | 0.8  (0.78 - 0.82) |
| Panic Disorder or Agoraphobia | >52 | 87.8%  (73.8% - 95.9%) | | 65.96%  (63.3% - 68.5%) | | 0.54%  (0.39% - 0.62%) | 13.4  (11.9 - 15.1) | | 98.9  (97.5 - 99.5) | | 2.58  (2.25 - 2.96) | | 0.18  (0.08 - 0.42) | | 0.82  (0.79 - 0.84) |
| Post-traumatic Stress Disorder | >57 | 58.82%  (32.9% - 81.6%) | | 83.51%  (81.4% - 85.4%) | | 0.42%  (0.24% - 0.61%) | 7.1  (4.8 - 10.4) | | 99.0  (98.2 - 99.4) | | 3.57  (2.35 - 5.4) | | 0.49  (0.28 - 0.87) | | 0.75  (0.73 - 0.77) |
| Attention Deficit Hyperactivity Disorder | >51 | 62.96%  (42.4% - 80.6%) | | 61.25%  (58.6% - 63.9%) | | 0.24%  (0.09% - 0.36%) | 3.8  (2.8 - 5) | | 98.6  (97.7 - 99.1) | | 1.62  (1.21 - 2.19) | | 0.60  (0.37 - 0.99) | | 0.63  (0.61 - 0.66) |
| Conduct Disorder | >57 | 55.17%  (35.7% - 73.6%) | | 83.81%  (81.7% - 85.7%) | | 0.39%  (0.2% - 0.53%) | 8.7  (6.3 - 11.9) | | 98.5  (97.8 - 99.0) | | 3.41  (2.4 - 4.83) | | 0.53  (0.36 - 0.8) | | 0.72  (0.7 - 0.75) |

**Note:** Optimal cut points were determined by the analysis of ROC (Receiver Operating Characteristic) curves, balancing sensitivity and specificity. **Abbreviations:** CI=Confidence Interval

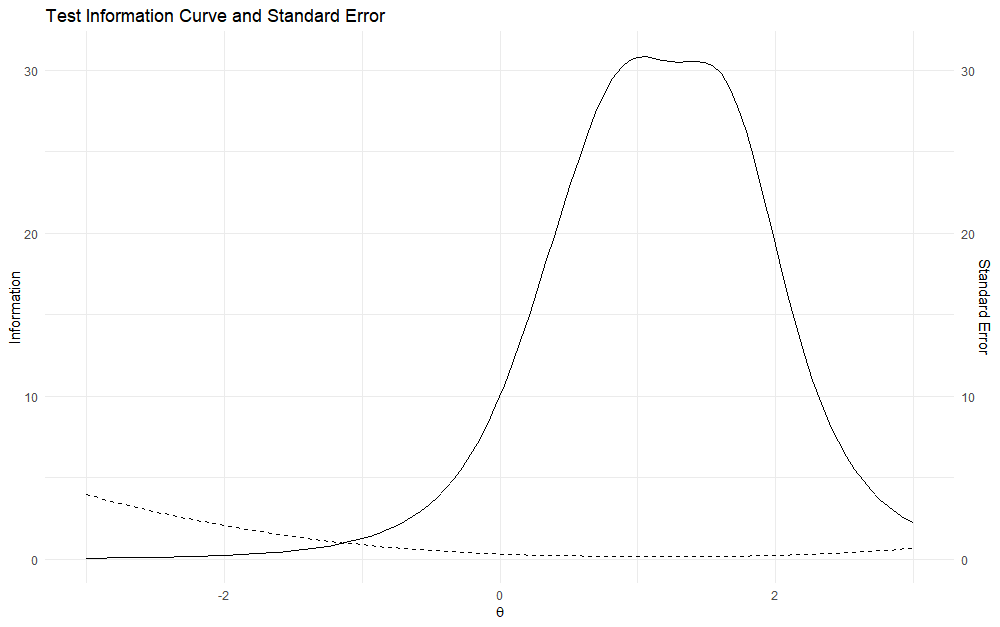

**Figure S1.** Test information curve (solid line) and standard error (dashed line) of the Short Mood and Feelings Questionnaire (SMFQ).

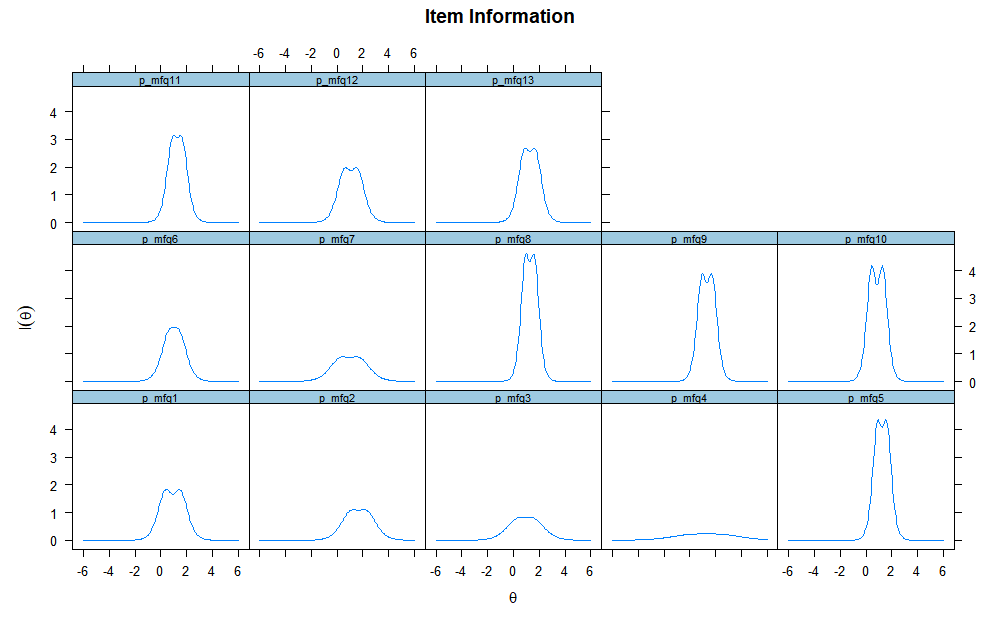

**Figure S2.** Item information curves for the 13 items of the Short Mood and Feelings Questionnaire (SMFQ).

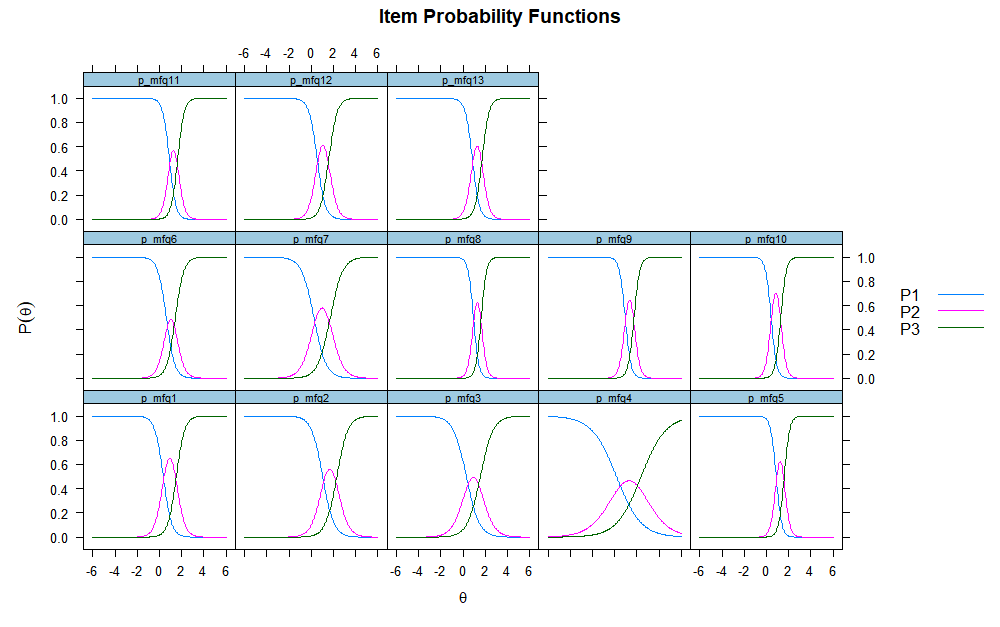

**Figure S3.** Item characteristic curves for the 13 items of the Short Mood and Feelings Questionnaire (SMFQ).

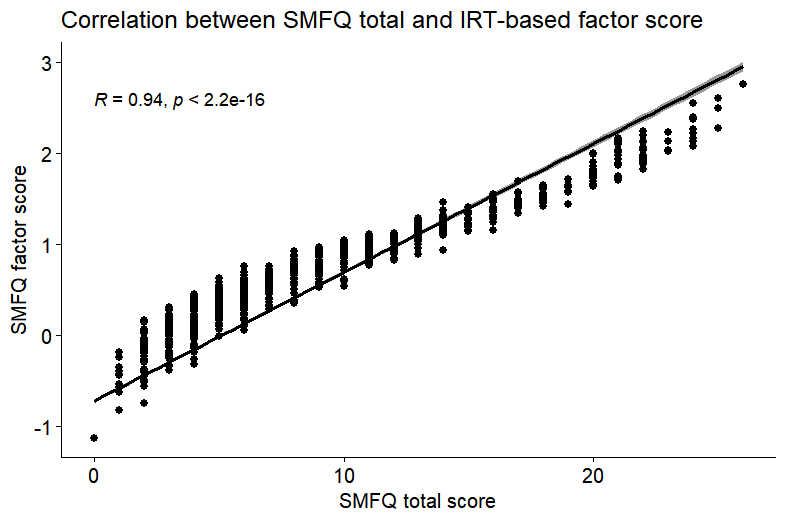

**Figure S4.** Correlation between Short Mood and Feelings Questionnaire (SMFQ) total and IRT-based factor score.

**Note:** SMFQ = Short Mood and Feelings Questionnaire.

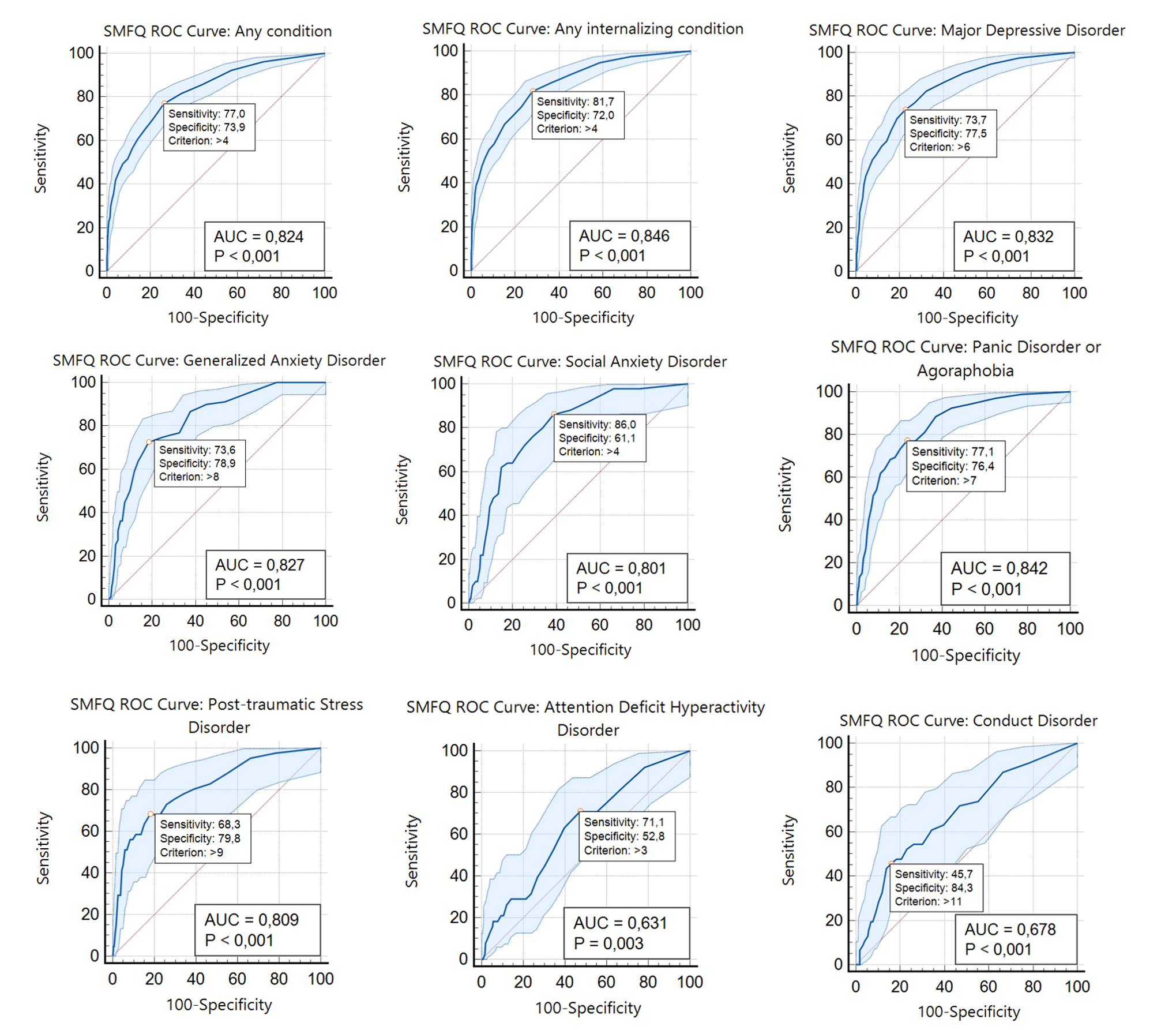

**Figure S5.** SMFQ (sum scores) ROC Curves for all mental health conditions with prevalence ≥ 2% in this sample.

**Note:** ROC (Receiver Operating Characteristic) were generated using the SMFQ sum score against DAWBA diagnosis. The highlighted point represents the chosen cut point for this sample and its sensitivity and specificity properties, respectively. ROC = Receiver Operating Characteristic. AUC = Area Under the Curve.

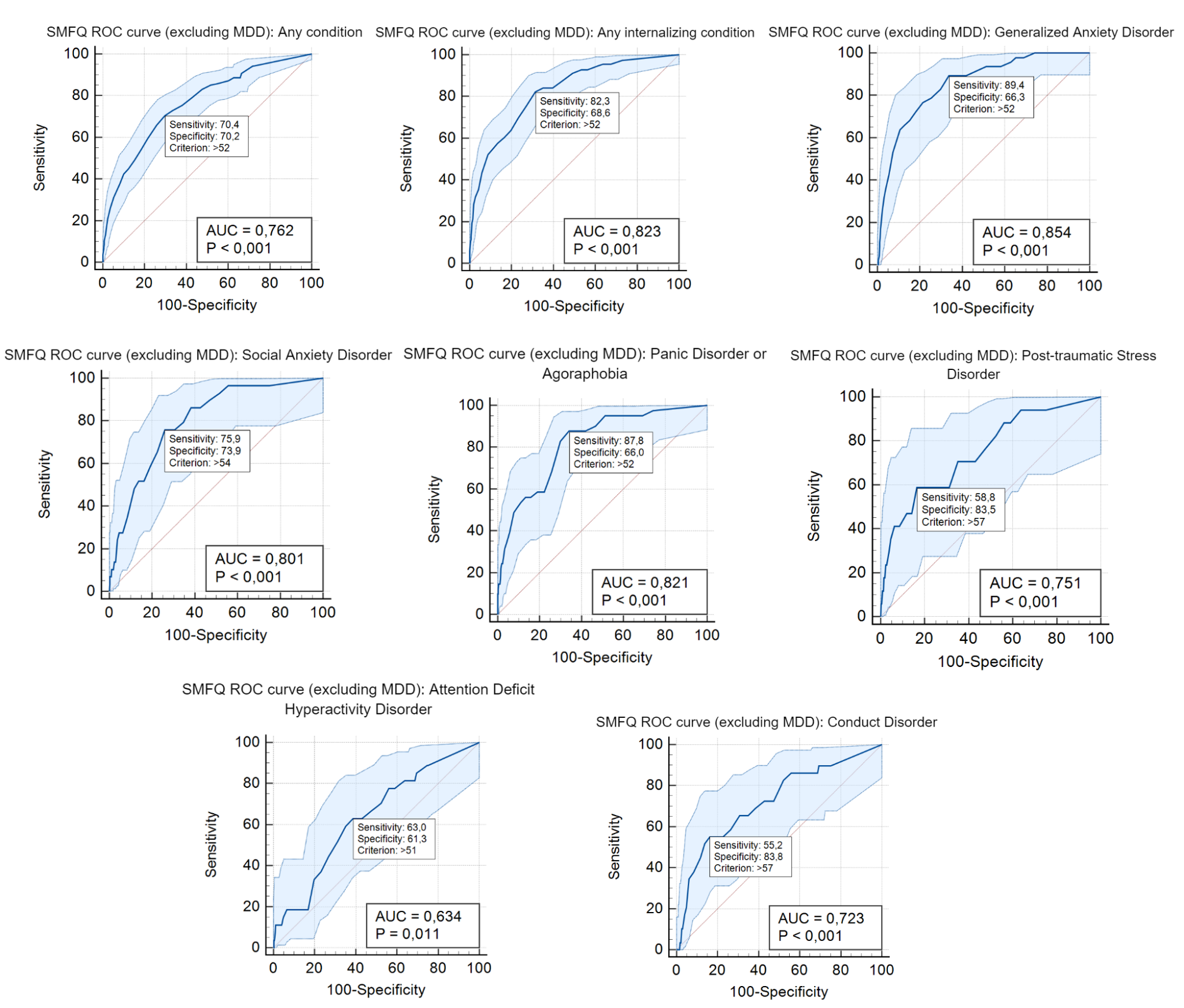

**Figure S6.** SMFQ (T-score) ROC Curves for all mental health conditions with prevalence ≥ 2% in this sample, excluding major depressive disorder diagnosis.

**Note:** ROC (Receiver Operating Characteristic) were generated using the SMFQ T-score against DAWBA diagnosis. The highlighted point represents the chosen cut point for this sample and its sensitivity and specificity properties, respectively. ROC = Receiver Operating Characteristic. AUC = Area Under the Curve.
